# The predictive value of heparin-binding protein for bacterial infections in patients with severe multiple trauma

**DOI:** 10.1101/2024.03.05.24303814

**Authors:** Li Li, Xiao-xi Tian, Gui-long Feng, Bing Chen

**Affiliations:** Department of Critical Care Medicine, The Second Hospital of Tianjin Medical University, Tianjin 300070, China; Department of Emergency, The First Hospital of Shanxi Medical University, Taiyuan 030001, Shanxi, China; Department of Emergency, The Second Hospital of Tianjin Medical University, Taiyuan 030001, Shanxi, China

**Keywords:** multiple trauma, heparin-binding protein, bacterial, infection

## Abstract

**Introduction:** Heparin-binding protein is an inflammatory factor with predictive value and participates in the inflammatory response through antibacterial effects, chemotaxis, and increased vascular permeability. The role of heparin-binding protein in sepsis has been progressively demonstrated, but few studies have been conducted in the context of multiple trauma combined with bacterial infections. This study aims to investigate the predictive value of heparin-binding protein for bacterial infections in patients with severe multiple trauma.

**Materials and methods:** Patients with multiple trauma in the emergency intensive care unit were selected for the study, and plasma heparin-binding protein concentrations and other laboratory parameters were measured within 48 hours of admission to the hospital. A two-sample comparison and univariate logistic regression analysis were used to investigate the relationship between heparin-binding protein and bacterial infection in multiple trauma patients. A multifactor logistic regression model was constructed, and the ROC curve was plotted.

**Results:** Ninety-seven patients with multiple-trauma were included in the study, 43 with bacterial infection and 54 without infection. According to data analysis, heparin-binding protein was higher in the infected group than in the control group [(32.00±3.20) ng/mL *vs.* (18.52±1.33) ng/mL]. Univariate logistic regression analysis shows that heparin-binding protein is related to bacterial infection (*OR*=1.10, *Z*=3.91, *95%CI*:1.05∼1.15, *P*=0.001). Multivariate logistic regression equations showed that patients were 1.12 times more likely to have bacterial infections for each value of heparin-binding protein increase, holding neutrophils and PCT constant. ROC analysis shows that heparin-binding protein combined with neutrophils and PCT has better predictive value for bacterial infection [*AUC*=0.935, *95%CI*:0.870∼0.977].

**Conclusions:** Heparin-binding protein may predict bacterial infection in patients with severe multiple trauma. Combining heparin-binding protein, PCT, and neutrophils may improve bacterial infection prediction.

## Introduction

Multiple trauma is the fourth leading cause of death, accounting for 11.2% of disability-adjusted life-years worldwide [1,2]. Infection and inflammatory response after multiple trauma greatly impact the prognosis. Therefore, monitoring inflammatory indicators can help early identification of infection and timely adjustment of treatment strategies [3]. Although the Advanced Trauma Life Support clinical guideline and the Injury Severity Score can assess the condition and guide interventions, more reliable indicators of infection are needed for early prediction of infection [4]. Given the critical role of inflammatory factors in multiple injuries [5], using reliable indicators to predict infection is essential for early clinical intervention and reduction of mortality.

Heparin-binding protein (HBP) is mainly produced and stored in neutrophils [6]. It is released in response to numerous cytokines, inflammatory agents, chemokines, and microorganisms. Monocytes can also release small amounts of it [7,8]. HBP possesses chemotactic and antimicrobial activities, participates in inflammatory responses, and can promote vascular permeability. It is also known as Azurocidin or 37,000 cationic antimicrobial protein (CAP37) [9]. The diagnostic and prognostic predictive role of heparin-binding protein in sepsis has been progressively demonstrated, but few studies have been conducted in the context of multiple trauma combined with bacterial infections [10-12].

The purpose of this study was to determine whether HBP predicts bacterial infection in patients with severe multiple trauma. For this purpose, we prospectively studied the correlation between HBP and bacterial infection and selected variables with clinical value to construct a logistic regression model. The calibration curve and ROC curve are used to evaluate the accuracy and predictive value of the model, respectively. It provides a reference value for predicting bacterial infections in emergency multiple trauma in clinical practice.

## Methods

### Sample sources and ethical issues

This is a prospective study of patients with multiple-trauma admitted to the Emergency Intensive Care Unit of the First Hospital of Shanxi Medical University from November 2022 to October 2023. All procedures were performed in accordance with relevant laws and institutional guidelines, in compliance with the World Medical Association Declaration of Helsinki, and were approved by the Ethics Committee of the First Hospital of Shanxi Medical University [2021 Lun Review Words (K-K115)]. Case details and personal information included in this study were obtained with consent and authorization. Written consent for this study was obtained from the participants involved or from the patients’ families.

The outcome indicators were the patients’ blood, cerebrospinal fluid, and sputum culture findings. Patients with no positive microbiologic culture findings were placed in the control group, while those with positive bacteria culture results were placed in the infection group.

### Inclusion and exclusion criteria

The inclusion criteria included the following: Age ≥18 years; time from injury to admission <24 hours; patients diagnosed with severe multiple trauma; first hospitalization for multiple trauma. Exclusion Criteria: Patients with a hospital stay of less than three days or who are certified dead upon resuscitation; patients with other infections that are not bacterial; prior use of anticoagulant medications; infectious disease before the current trauma; history of severe trauma or surgery in the last three months; diseases that affect coagulation or immunity.

### Sample detection and data collection

Within 48 hours of the patient’s admission, peripheral venous or preoperative blood collection was performed. Avoiding blood collection with heparin-containing indwelling needles. A 5 ml tube containing sodium citrate was used to collect blood. Samples were centrifuged within 5 minutes of blood collection (5200 r/min, 5 min). After centrifugation, the plasma was taken 20μl) and refrigerated at 2∼8°C for five days. The HBP assay was carried out in batches using rate-scattering turbidimetric assay to reduce the error (The reagents and instruments used to detect HBP in this study were obtained from Suzhou Kangheshun Medical Co.). Patient clinical information is based on information available during this admission. Laboratory indicators are based on the return of laboratory results within 48 hours of our hospital.

### Statistical analysis

Stata MP 16 was used to carry out all statistical evaluations and visualizations. Quantitative was described as mean ± standard deviation, and qualitative was described using counts and percentages. Quantitative comparisons were made using the two independent samples t-test or *t-test*. Qualitative comparisons were made using the *χ2* or *Fisher’s exact test*. *P*<0.05 was considered statistically significant.

Univariate variable logistic analyses were performed on variables whose differences were statistically significant after comparing the two samples. The multivariate logistic regression model was constructed using forward stepwise regression to screen the significant variables after univariate logistic regression analysis. Hosmer-Lemeshow test was used to test the goodness of fit of the model. The calibration curve and ROC curve were used to visualize and evaluate the predictive efficacy of the logistic regression model.

## Result

### Clinical characteristics and comparison of two groups

After screening and testing, 97 multiple-trauma patients were eligible and participated in this study. Microbiological cultures were performed based on the patient’s blood, sputum, and cerebrospinal fluid, and 43 were eventually included in the bacterial infection group and 54 in the control group. The clinical characteristics and comparison of the two groups are shown in Table 1. HBP was higher in the infected group than in the control group, and the difference was statistically significant [(32.00±3.20) ng/mL *vs.* (18.52±1.33) ng/mL]. Besides, Age, smoking, respiratory rate, HBP, leukocytes, neutrophils, and PCT were higher in the infected group than in the control group, and the difference was statistically significant(*P*<0.05). The control group had a higher GCS score than the infected group, and the difference was statistically significant(*P*<0.05). Gender, BMI, temperature, pulse, ISS score, alcohol consumption, hypertension, diabetes mellitus, lymphocytes, calcium, erythrocytes, hemoglobin, platelets, PT, PTINR, APTT, TT, FDP, DD, FIBc had no statistically significant difference between the two groups(*P*>0.05).

**Table 1.** Clinical characteristics and comparison of two groups.

|  | Infection group<br>(*n*=43) | Control group<br>(*n*=54) | Overall<br>(*n*=97) | *P* |
| --- | --- | --- | --- | --- |
| **Gender, *n* (%)** |  |  |  | 0.346 |
| Female | 7 (16.28) | 13 (24.07) | 20 (20.62) |  |
| Male | 36 (83.72) | 41 (75.93) | 77 (79.38) |  |
| **Age, years** | 58.88±2.40 | 51.80±1.98 | 54.94±1.56 | **0.008** |
| **BMI (kg/m<sup>2</sup>)** | 23.44±0.39 | 24.49±0.49 | 24.02±0.33 | 0.095 |
| **Body temperature** | 36.67±0.08 | 36.60±0.07 | 36.63±0.05 | 0.854 |
| **Pulse** | 83.12±2.94 | 81.76±2.12 | 82.36±1.75 | 0.931 |
| **Respiration** | 19.65±0.63 | 18.15±0.56 | 18.81±0.42 | **0.019** |
| **ISS** | 20.42±1.19 | 18.28±1.17 | 19.23±0.84 | 0.094 |
| **GCS** | 8.72±0.57 | 11.20±0.41 | 10.10±0.16 | **0.001** |
| **Tobacco Smoking, *n* (%)** | 27 (62.79) | 20 (37.04) | 47 (48.45) | **0.012** |
| **Alcohol Drinking, *n* (%)** | 18 (41.86) | 13 (24.07) | 31 (31.96) | 0.062 |
| **Comorbidities, *n* (%)** |  |  |  |  |
| Hypertension | 24 (55.81) | 22 (40.74) | 46 (47.42) | 0.140 |
| Diabetes | 8 (18.60) | 5 (9.26) | 13 (13.40) | 0.180 |
| **Peripheral blood** |  |  |  |  |
| HBP (ng/mL) | 32.00±3.20 | 18.52±1.33 | 24.5±1.73 | **0.001** |
| WBC (10<sup>9</sup>/L) | 14.49±0.79 | 8.96±0.50 | 11.41±0.53 | **0.001** |
| Lymphocyte (10<sup>9</sup>/L) | 1.41±0.39 | 1.45±0.23 | 1.43±0.21 | 0.156 |
| Neutrophil (10<sup>9</sup>/L) | 12.36±0.75 | 6.84±0.51 | 9.29±0.52 | **0.001** |
| PCT (10<sup>9</sup>/L) | 1.10±0.17 | 0.27±0.04 | 0.64±0.09 | **0.001** |
| Ca (mg/dL) | 2.20±0.02 | 2.23±0.02 | 2.22±0.01 | 0.383 |
| RBC (10<sup>12</sup>/L) | 4.07±0.21 | 4.12±0.11 | 4.1±0.11 | 0.287 |
| HB (g/L) | 121.20±3.43 | 123.32±3.39 | 122.38±2.41 | 0.703 |
| PLT (10<sup>9</sup>/L) | 197.67±14.86 | 193.35±7.80 | 195.26±7.85 | 0.470 |
| PT (s) | 14.80±0.43 | 14.65±0.17 | 14.72±0.21 | 0.850 |
| PTINR | 1.19±0.10 | 1.14±0.08 | 1.16±0.06 | 0.569 |
| APTT (s) | 27.24±0.57 | 26.76±0.38 | 26.97±0.33 | 0.700 |
| TT (s) | 15.51±0.34 | 15.55±0.31 | 15.53±0.23 | 0.862 |
| FDP (ug/ml) | 27.73±3.55 | 21.70±2.34 | 24.37±2.05 | 0.309 |
| DD (ng/ml) | 2113.28±311.89 | 1805.07±239.24 | 1941.70±191.58 | 0.452 |
| FIB c (g/L) | 3.40±0.17 | 3.10±0.10 | 3.23±0.10 | 0.110 |
| **Surgery, *n* (%)** | 31 (72.09) | 29 (53.70) | 60 (61.86) | 0.064 |

Data are presented as numbers, percentages, and mean ± standard deviation; ISS, Injury Severity Score;
GCS, Glasgow Coma Scale; HBP, heparin-binding protein; WBC, white blood cell; PCT, procalcitonin;
Ca, Calcium; RBC, red blood cell; HB, hemoglobin; PLT, blood platelet; PT, prothrombin time; PTINR,
prothrombin time and international normalized ratio; APTT, activated partial thromboplastin time; TT,
thrombin time; FDP, fibrinogen degradation product; DD, D dimer; FIBc, fibrinogen.

A total of 43 patients were identified in the bacterial infection group by patient blood, sputum, and cerebrospinal fluid cultures. Bacterial species and distribution are shown in Table 2. Among the patients with positive bacterial cultures, eight patients had multiple bacterial infections. More patients had positive sputum cultures for bacteria (*n*=39) than cerebrospinal fluid cultures (*n*=8), and fewer patients had positive blood cultures for bacteria (*n*=4). In addition, the most common result for sputum cultures that were positive for bacteria was Klebsiella pneumoniae (*n*=16), and the most common bacteria for cerebrospinal fluid infections was Staphylococcus (*n*=5). Bacteria positive in blood culture include Staphylococcus, Enterobacteriaceae, Fungal, and Bacillus.

**Table 2.** Bacterial species and distribution.

| Bacterial species | Sputum culture | Blood Culture | Cerebrospinal Fluid |
| --- | --- | --- | --- |
| *Staphylococcus* ($n$) | 4 | 1 | 5 |
| *Klebsiella pneumoniae* ($n$) | 16 | 0 | 1 |
| *Enterobacteriaceae* ($n$) | 7 | 1 | 1 |
| *Stenotrophomonas* ($n$) | 1 | 0 | 0 |
| *Moraxellaceae* ($n$) | 4 | 0 | 0 |
| *Fungal* ($n$) | 1 | 1 | 1 |
| *Haemophilus* ($n$) | 1 | 0 | 0 |
| *Corynebacterium striatum* ($n$) | 5 | 0 | 0 |
| *Bacillus* ($n$) | 0 | 1 | 0 |
| Count ($n$) | 39 | 4 | 8 |

### Univariate logistic regression analysis

Univariate logistic regression was used to analyze the variables with statistically significant differences between the infection and control groups, as shown in Table 3. The results show that age, GCS score, smoking, HBP, WBC, neutrophils, and PCT are related to bacterial infection in multiple trauma (all *P*<0.05). Besides, Respiration has no significant correlation with multiple-trauma infection (*OR*=1.10, *Z*=1.70, *95%CI*:0.99∼1.22, *P*=0.09). Furthermore, HBP is associated with bacterial infections in multiple trauma (*OR*=1.10, *Z*=3.91, *95%CI*:1.05∼1.15, *P*=0.001). This confirms that the HBP difference between the infected and control groups was statistically significant.

**Table 3.** Univariate logistic regression analysis.

|  | *Odds ratio* | *Standard Error* | *Z* | *P>Z* | *[95% conf. interval]* |  |
| --- | --- | --- | --- | --- | --- | --- |
| Age, years | 1.03 | 0.02 | 2.19 | 0.028 | 1.00 | 1.06 |
| Respiration | 1.10 | 0.06 | 1.70 | 0.090 | 0.99 | 1.22 |
| GCS | 0.81 | 0.05 | -3.27 | 0.001 | 0.71 | 0.92 |
| Tobacco Smoking, *n* (%) | 2.87 | 1.21 | 2.49 | 0.013 | 1.25 | 6.57 |
| HBP (ng/mL) | 1.10 | 0.03 | 3.91 | 0.001 | 1.05 | 1.15 |
| WBC ($10^9/L$) | 1.44 | 1.12 | 4.34 | 0.001 | 1.22 | 1.70 |
| Neutrophil ($10^9/L$) | 1.44 | 0.12 | 4.41 | 0.001 | 1.22 | 1.69 |
| PCT ($10^9/L$) | 24.47 | 17.92 | 4.37 | 0.001 | 5.83 | 102.78 |

GCS, Glasgow Coma Scale; HBP, heparin-binding protein; WBC, white blood cell; PCT, procalcitonin.

### Multivariable logistic regression model

In order to further screen out variables with statistical significance and clinical value, the variables after univariate logistic analysis were screened using forward stepwise regression, and a multivariate logistic regression model was obtained, as shown in Table 4. The variables screened out by forward stepwise regression included HBP, neutrophils, and PCT. Multivariate logistic regression equations showed that patients were 1.12 times more likely to have bacterial infections for each value of heparin-binding protein increase, holding neutrophils and PCT constant.

**Table 4.** Multivariable logistic regression model.

|  | *Odds ratio* | *Standard Error* | *Z* | *P* | *[95% conf. interval]* |  |
| --- | --- | --- | --- | --- | --- | --- |
| **HBP** | 1.118 | 0.042 | 2.95 | 0.003 | 1.038 | 1.205 |
| **Neutrophil** | 1.204 | 0.090 | 2.47 | 0.013 | 1.039 | 1.395 |
| **PCT** | 53.876 | 57.207 | 3.75 | 0.001 | 6.723 | 431.735 |
| **constant** | 0.001 | 0.002 | -4.69 | 0.001 | 0.001 | 0.021 |

HBP, heparin-binding protein; PCT, procalcitonin.

### Hosmer-Lemeshow test and Calibration curve

The *Hosmer-Lemeshow test* (*HL test*) was used to analyze the goodness of fit of the multivariable logistic regression model. The results show that the *P* value is insignificant, so the PH assumption cannot be violated (*χ^2^*=86.96, *P*= 0.6568>0.05). The model passes the *HL test,* and the model fit is good. The calibration curve is plotted to visualize the logistic regression model based on the results of the *HL* goodness-of-fit test, as shown in Figure 1. The deviation between predicted and observed risk is slight and close to the reference line, indicating that the model’s predictive ability is good.

**Figure 1.**
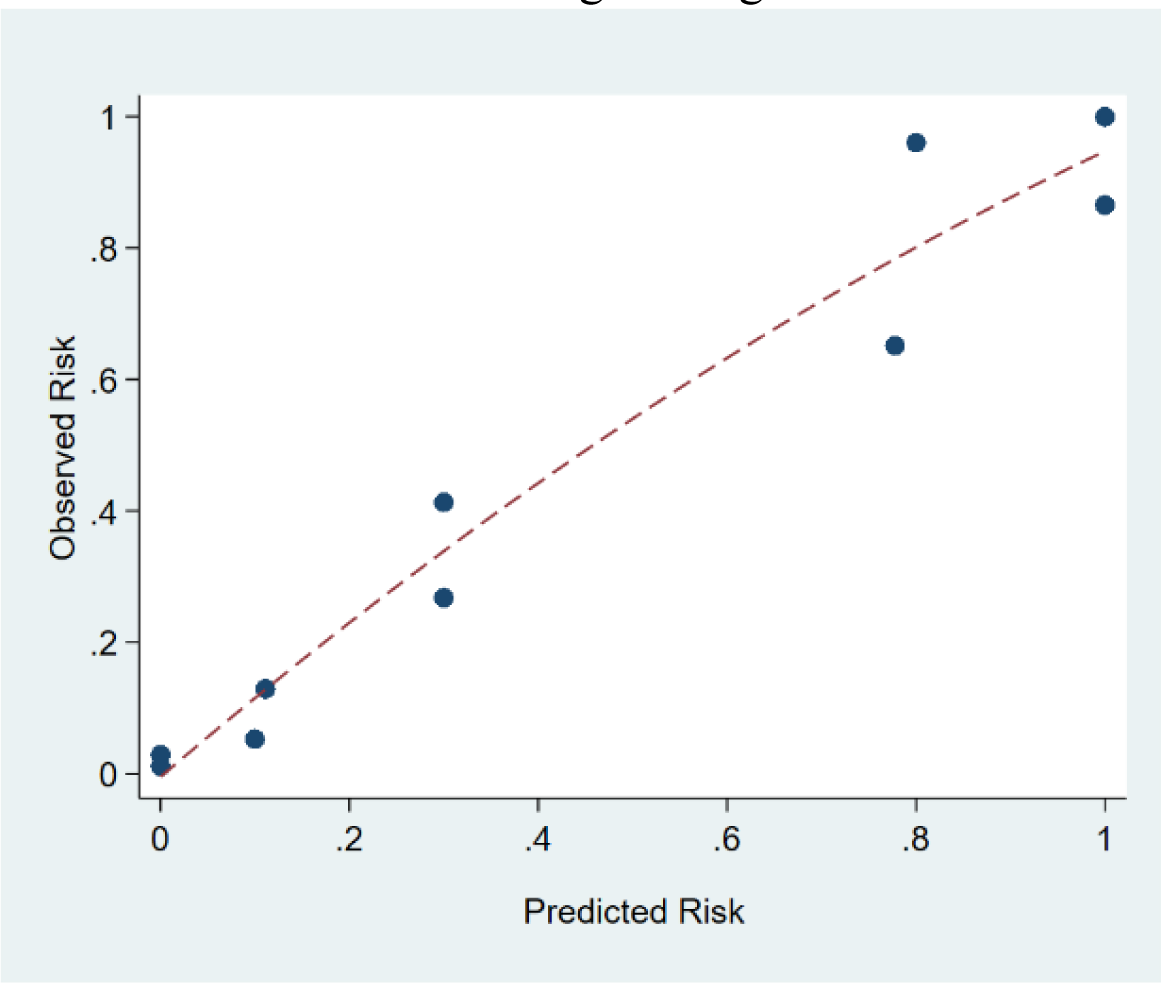
Calibration curve of multivariable logistic regression model.

### ROC analysis

ROC analysis was used to assess the predictive effectiveness of HBP and multivariate logistic regression models. The results showed that the Area Under Curve (*AUC*) for HBP alone to predict infection in multiple trauma was 0.773 (the optimal cut-off value of HBP was 18.08, with a sensitivity of 86.05%, a specificity of 55.56%, and a Yordon index of 0.416 (*AUC*=0.773, *95% CI*: 0.677∼0.852). ROC curves were plotted for the multivariate logistic regression model for comparison, and the results showed that the multivariate logistic regression model had an AUC of 0.935 (*95% CI*:0.870∼0.977), and its predictive efficacy was better than that of HBP alone, as shown in Figure 2.

**Figure 2.**
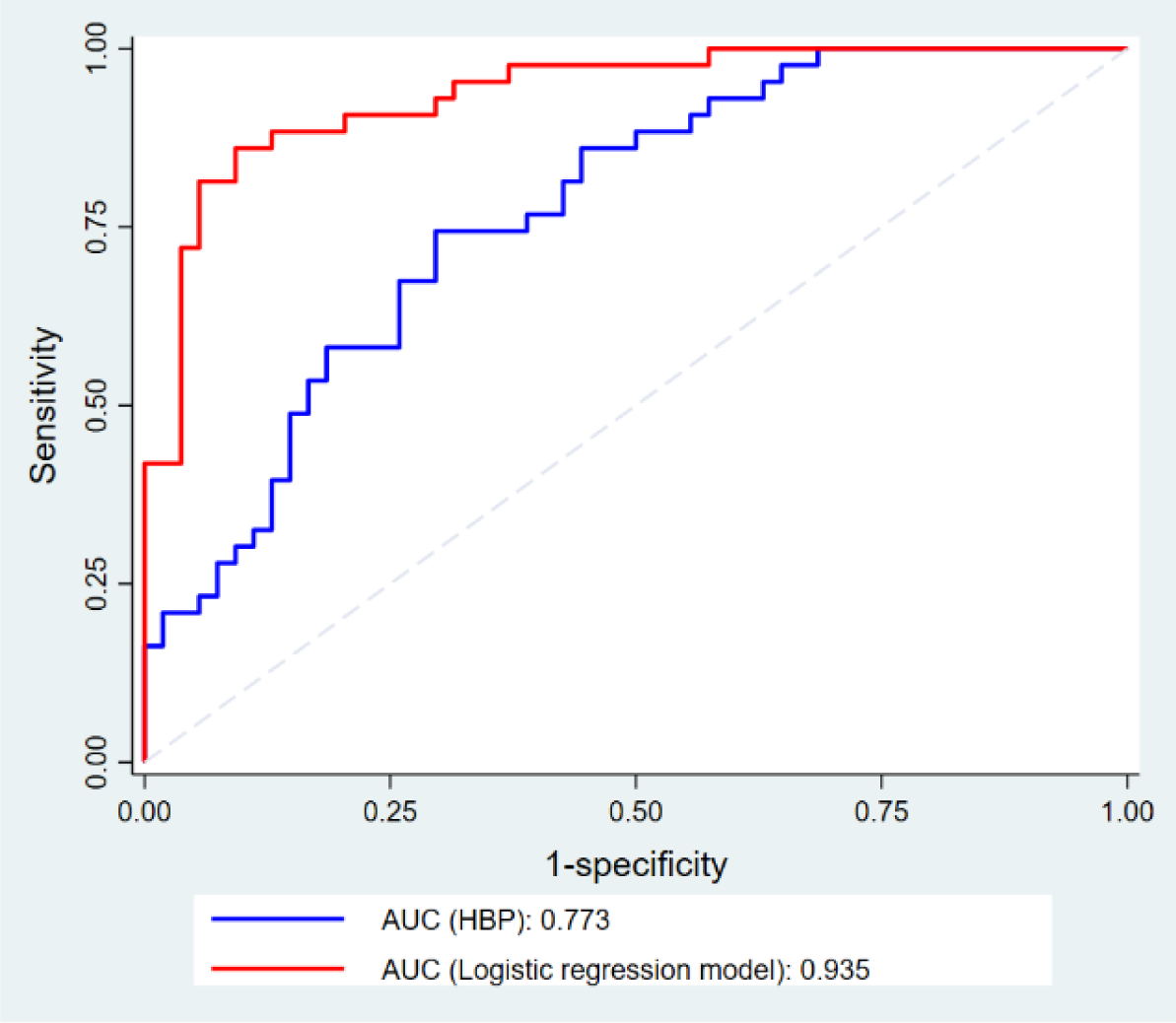
ROC curves for heparin-binding protein and multivariable logistic regression models.

## Discussion

To clarify the predictive value of HBP for bacterial infections in multiple trauma, we utilized the parameter test and univariate logistic regression to analyze the correlation between HBP and infection in multiple trauma. The forward stepwise regression was utilized to screen for clinically valuable variables, a multivariate logistic regression model was constructed, and the model was tested. Our study shows that HBP is a potential predictor of bacterial infection in patients with severe multiple trauma. Combining HBP, PCT, and neutrophils may improve the prediction of bacterial infection.

In this study, we found that HBP concentrations were higher in the infected group than in the control group, indicating a link between HBP and bacterial infections. Several studies in related fields demonstrated that HBP has antibacterial activity against gram-negative and gram-positive bacteria [13–15]. As shown in the research of H. Anne Pereira et al., the antibacterial activity of HBP may be related to its amino acids 20-44 acting as antibacterial domains [16].

Similarly, in univariate logistic regression analysis, we found that HBP correlated with bacterial infection in multiple trauma, confirming the previous parameter test results. The reason for this may be the involvement of HBP in the early immune response to bacterial infection in polytrauma [17–18]. In addition, univariate logistic regression analysis showed that age, GCS, Tobacco Smoking, HBP, WBC, Neutrophil, and PCT were all associated with bacterial infection (*P*<0.05). On the one hand, the results of univariate analyses can be affected by confounding factors, so univariate analyses are primarily used to make preliminary judgments about which factors are associated with infection. On the other hand, the analytical results of single-centre studies can be regionally biased. Therefore, we further screened for statistically significant variables in univariate logistic regression analyses.

To clarify the predictive value of HBP for bacterial infections in patients with multiple trauma, we performed multifactor logistic regression analysis using forward stepwise regression. Variables that were statistically significant and clinically valuable after screening were included in the optimal regression equation, including HBP, neutrophils, and PCT. Multivariate logistic regression equations showed that HBP, in combination with other indicators of inflammation, had a better predictive value for predicting bacterial infections. The HL test and Calibration curve results show that the multivariable logistic regression model has good fit and predictive ability. Our findings on the predictive value of HBP combined with other inflammatory indicators agree with those reported by Jin Ma et al., who discovered that adding HBP to other inflammatory markers enhanced prediction accuracy [19]. Furthermore, ROC analysis confirmed that the predictive value of the logistic regression model was superior to that of HBP alone, showing that HBP can be used as a better complementary indicator to existing inflammatory indicators.

The role of HBP as an inflammatory factor in infectious diseases has been progressively identified and confirmed [20]. For the first time, we showed the predictive value of HBP for bacterial infections in patients with multiple trauma. HBP has good predictive value for bacterial infections in patients with multiple trauma patients in the emergency department, and HBP combined with neutrophils and PCT better predicts bacterial infections. Therefore, HBP provides a better laboratory basis for early assessment of the condition of trauma patients. However, our study had several limitations. First, as this was a single-centre study, multicentre studies with larger sample sizes are needed to validate HBP’s predictive value further. Second, the condition of patients with multiple-trauma in emergency medicine is urgent and complex, so we could not classify and analyze them according to the injury site. Third, the role of HBP in non-bacterial infections such as viruses, parasites, and fungi needs to be investigated and validated. The next step is to optimize experimental plans and expand experimental samples.

## Conclusions

In patients with severe multiple trauma, heparin-binding protein may predict bacterial infection. Combining heparin-binding protein, PCT, and neutrophils may improve bacterial infection prediction. HBP provides a referenceable test for early disease assessment.

## CRediT authorship contribution statement

**Li Li:** Conceptualization, Data curation, Investigation, Methodology, Software, Visualization, Writing-original draft. **Xiao-xi Tian:** Data curation, Data curation, Investigation, Methodology. **Gui-long Feng:** Conceptualization, Formal analysis, Project administration, Resources, Writing-review. **Bing Chen:** Conceptualization, Formal analysis, Project administration, Resources, Supervision, Writing-editing. Bing Chen and Gui-long Feng contributed equally to this work and should be considered co-corresponding authors.

## Ethical approval

The study complied with the Declaration of Helsinki of the World Medical Association. This study was approved by the Ethics Committee of the First Hospital of Shanxi Medical University [2021 伦审字(K-K115)]. This article does not contain any studies with animals performed by any authors. The study complied with the Declaration of Helsinki of the World Medical Association. All participants or family members were aware of the risks and signed an informed consent form. This article does not contain any studies with animals performed by any authors.

## Data Availability

This text is appropriate if the data are owned by a third party and authors do not have permission to share the data.

